# Modeling COVID-19 vaccine efficacy and coverage towards herd-immunity in the Basque Country, Spain

**DOI:** 10.1101/2021.07.12.21260390

**Authors:** Nico Stollenwerk, Javier Mar, Joseba Bidaurrazaga Van-Dierdonck, Oliver Ibarrondo, Carlo Estadilla, Maíra Aguiar

## Abstract

Vaccines have measurable efficacies, obtained first from vaccine trials. However, vaccine efficacy is not a static measure upon licensing, and the long term population studies are very important to evaluate vaccine performance and impact. COVID-19 vaccines were developed in record time and although the extent of sterilizing immunity is still under evaluation, the currently licensed vaccines are extremely effective against severe disease, with vaccine efficacy significantly higher after the full immunization schedule. We investigate the impact of vaccines which have different efficacies after first dose and after the second dose administration schedule, eventually considering different efficacies against severe disease as opposed to overall infection. As a proof of concept, we model the vaccine performance of hospitalization reduction at the momentary scenario of the Basque Country, Spain, with population in a mixed vaccination setting, giving insights into the population coverage needed to achieve herd immunity in the current vaccination context.

## 1. Introduction

More than a year has passed since a severe respiratory syndrome (COVID-19) caused by a new coronavirus (SARS-CoV-2) was identified in China [1]. Declared a pandemic by the World Health Organization (WHO) in March 2020 [2], COVID-19 has spread rapidly around the globe. As of July 1st, 2021, approximately 180 million cases were confirmed with more than 3.9 million deaths and a global case fatality ratio (CFR) of approximately 2% [3, 4].

With eventually substantial global underestimation of SARS-CoV-2 infection, COVID-19 symptoms can range from asymptomatic/mild to severe illness, and disease severity and death occurring according to a hierarchy of risks [5], with age and pre-existing health conditions enhancing disease severity.

Vaccines against COVID-19 have been developed in record time [6, 7, 8, 9]. With different efficacies, COVID-19 vaccines are remarkably effective against severe disease, however, the so called sterilizing immunity, occurring when vaccinated individuals cannot transmit the virus, is still being evaluated [10, 11]. Moreover, vaccine performance is expected to be driven by the ability of undetected asymptomatic infections transmitting the virus [12, 13], and therefore, a well planned strategy to use different COVID-19 vaccines will optimize hospitalization reductions.

Mathematical models convey ideas about the components of a host-pathogen interactions and have been intensively used to model the dynamical spreading of COVID-19. Acting as a tool to understand and predict the spread of the disease as well as to evaluate the impact of control in different epidemiological scenarios, several task forces were created to assist public health managers and governments during the COVID-19 pandemic.

Already in March 2020, a multidisciplinary task force (so-called Basque Modelling Task Force, BMTF) was created to assist the Basque health managers and the Basque Government during the COVID-19 responses. As an extension of the simple SIR model, a stochastic SHARUCD modeling framework was developed [14, 15, 16, 17, 18, 19]. Able to describe the COVID-19 epidemic in terms of disease spreading and control, and giving accurate projections on hospitalizations, ICU admissions and deceased cases, this framework is currently used to monitor the COVID-19 epidemic as lockdowns are relaxed and tightened.

In this paper, we investigate the impact of COVID-19 vaccination roll out in the Basque Country, Spain, using a flexible modeling framework taking into account the differences on a single dose and two dose immunization schedule over time. Studies like the one described here are of major importance to understand the impact of uneven roll out of vaccination, giving insights into future planning of immunization programmes of new vaccine generations that will need to be evaluated under the same settings presented here.

The paper is structured as follows. In Section 2 we describe the SHARUCD modeling framework currently used to assist the public health managers in the Basque Country during the COVID-19 crisis. Section 3 describes the baseline modelling framework used for the current model extensions. Section 4 presents the analytical results and numerical experiments with vaccine model using the latest data available on vaccine efficacy and coverage. The last section concludes this work with a discussion presenting the connection of this research with the current vaccination strategy to control COVID-19 spreading and severe disease reduction in the Basque Country, Spain.

## 2. Modeling COVID-19 in the Basque Country, Spain

As an extension of the simple SIR and SHAR models, the SHARUCD modeling framework considers populations of susceptible individuals (S), severe cases prone to hospitalization (H), mild, sub-clinical or asymptomatic (A), recovered (R), patients admitted to the intensive care units, ICU, (U). The recorded cumulative positive cases, which includes all new positive cases for each class of H, A, U, R, are counted within the C classes, including the deceased (D) cases [14, 15, 16, 17, 18].

Able to describe the COVID-19 epidemic in terms of disease spreading, the SHARUCD model gives accurate projections on hospitalizations, ICU admissions and deceased cases, from March 4, 2020 to December 2020, shown in Fig. 1, when vaccination started. The modeling framework was used to monitor the COVID-19 epidemiological dynamics in the Basque Country while the lockdown measures were relaxed and tightened over time.

**Figure 1:**
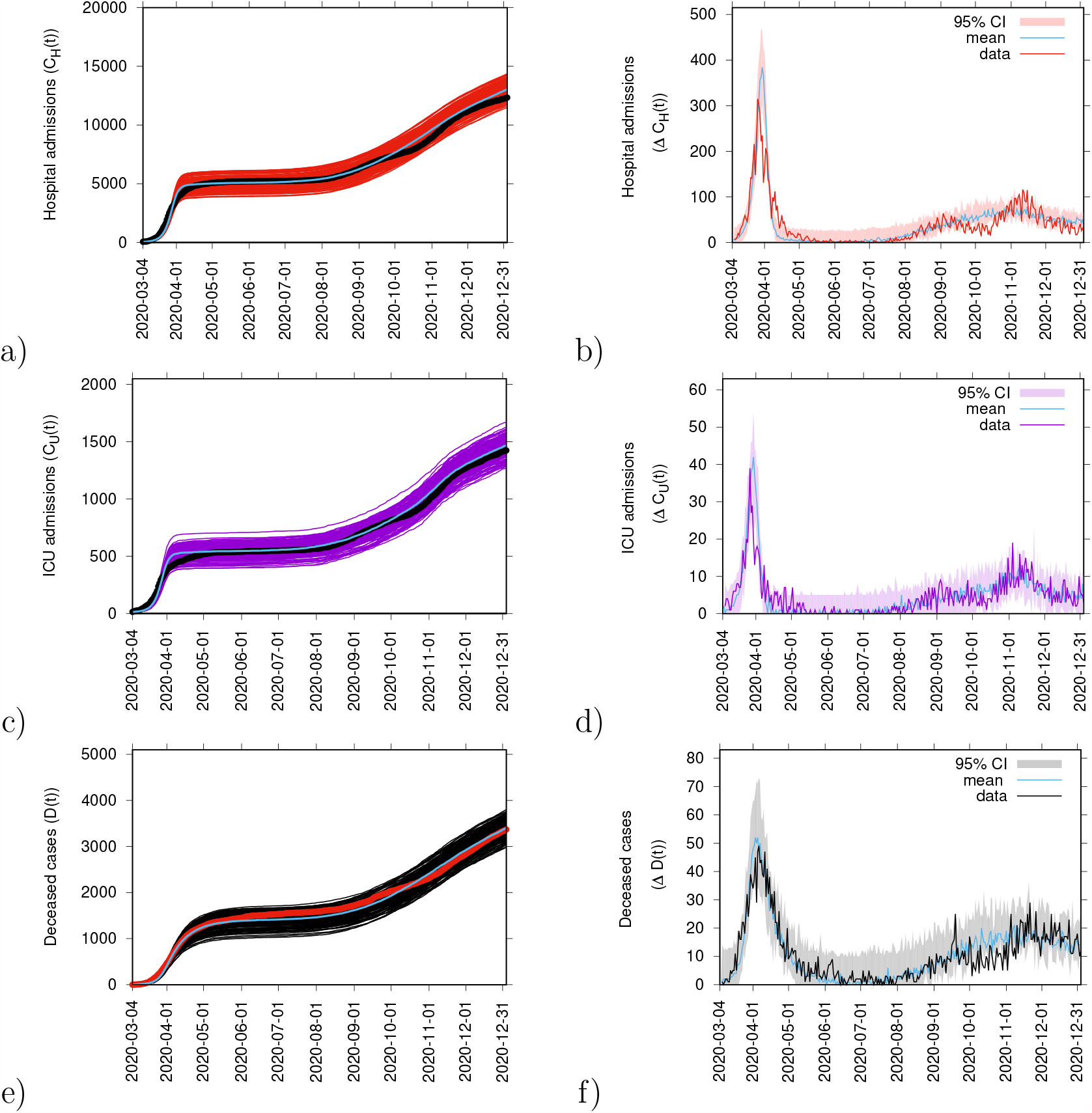
From March 4 to December 31, 2020, on the left hand side we plot the ensemble of stochastic realizations of the SHARUCD-model for cumulative cases. In a) cumulative hospitalized cases CH(t), in c) cumulative ICU admissions CU(t) and in e) cumulative deceases cases D(t). The mean the of the stochastic realizations is plotted in light blue. Empirical data are plotted for hospitalizations and for ICU admissions (black dots) and deceased cases (red dots). On the right hand side we plot the model results for the daily incidences. In b) daily hospitalized cases, in d) daily ICU admissions and in f) daily deceased cases. Empirical data are plotted for hospitalizations (red line), ICU admissions (purple line) and deceased cases (black line). The mean of 200 stochastic realizations are plotted as a light blue line. The 95% confidence intervals are obtained empirically from 200 stochastic realizations and are plotted as red, purple and black shadow for each disease related variable, hospitalizations, ICU admission and deceased cases, respectively.

However, to evaluate the implications of different vaccine efficacies and coverages, this framework is under refinement to include the uneven vaccination roll out strategy currently in place worldwide. As continuation of the BMTF efforts, we now evaluate the vaccination trial data for the vaccines which have been licensed for emergency use in Europe. Results are implemented into the simple SHAR modeling framework and its extensions to get the qualitative overview of the impact of COVID-19 vaccination strategy in the Basque Country and many other European regions.

### 2.1. Modeling COVID-19 vaccine trial and data analysis

Every vaccine must go through extensive and rigorous testing to ensure it is safe before it can be introduced in a country’s vaccine programme. The so called pre-clinical phase of a vaccine development determines which antigen invoke an immune response to a given pathogen. This phase is done without testing on humans. If the vaccine triggers an immune response, it will be then tested in human clinical trials which have three phases to assess its safety and confirm it generates an immune response. While the Phase I enrolls a small number of volunteers, in the Phase III vaccine is given to a much larger group of people, often across multiple countries, to determine the so called vaccine efficacy against the disease. For that, the studied population is divided into two groups: the vaccinated with individuals receiving the vaccine and the control group receiving a placebo solution [20]. These groups are compared to obtain the so called vaccine efficacy (VE) measure, i.e., the proportionate reduction in cases among vaccinated persons calculating the risk of disease among vaccinated and unvaccinated persons and determining the percentage of reduction in risk of disease among vaccinated persons relative to unvaccinated persons [21]. Vaccine efficacy is generally reported as relative risk (RR), the ratio of the probability of an outcome in an exposed group to the probability of an outcome in an unexposed group, which is calculated based on the number of confirmed infections, mild or severe, in each group. It is important to stress, however, that vaccine efficacy is not a static measure and therefore its evaluation continues further with eventually newly upcoming aspects of disease protection and preconditioning for its use being identified over time [22, 23, 24, 25].

In the present epidemiological scenario, COVID-19 were approved for emergency use according to current regulatory guidelines and legal requirements. Due to the global emergency, the vaccine trials lasted shorter time enrolling similar sample sizes for vaccine and placebo groups. As such, long term population studies and large scale field analysis, possible now after the global vaccination roll out, are very important to evaluate vaccine performance and impact.

Four vaccines are now used in the Basque Country, Spain, and other European countries. On the one hand, mRNA type vaccines BioNTech/Pfizer and Moderna, with above 90% vaccine efficacy estimated after second dose, while on the other viral vector vaccines by Oxford/AstraZeneca and Janssen/Johnson&-Johnson, with about 70% vaccine efficacy estimated upon full immunization schedule, with two or one dose respectively [6, 8, 9, 10].

The analysis of the raw vaccine trial data can be done by a Bayesian framework to estimate the efficacy conditioned on numbers of detected infected in the vaccine group and in the placebo group [26, 27, 28, 29].

In this section, we analyse the trial data from the Oxford/AstraZeneca vaccine [6], showing explicitly the COVID-19 vaccine efficacy as Bayesian posterior, and from this its cumulative distribution function, obtaining the confidence intervals in good agreement with reported numbers in [6].

Note that some vaccines have, by now, larger scale vaccine efficacy data publicly available. With a much larger group sizes, these studies are able to obtain information of various aspects of the vaccine efficacy after one dose versus full immunization, after the full immunization scheme of two doses, see e.g. for the BioNTech/Pfizer vaccine [10]. From such studies, the efficacy against severe disease/hospitalization and against infection can be obtained separately, for single dose and two dose regimes. Results of this analysis are shown in the section below, and will be also included into the modelling framework to evaluate the expected impact of vaccination campaigns in a populations with known vaccine coverage of individuals receiving two doses and for at least one dose.

### 2.2. Analysis of the Oxford/AstraZeneca COVID-19 vaccine trial

The Oxford/AstraZeneca vaccine trial in the UK/Brazil study [6] has roughly the same sample size for control group and vaccine group, i.e, *N*_*v*_ ≈ *N*_*c*_. While the control group *N*_*c*_ = 5829 reported *I*_*c*_ = 101 infected individuals, the vaccine group *N*_*v*_ = 5807 counted *I*_*v*_ = 30 infected individuals, giving a maximum likelihood estimate for the vaccine efficacy of 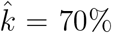, as also reported in [6]. This estimation was obtained via

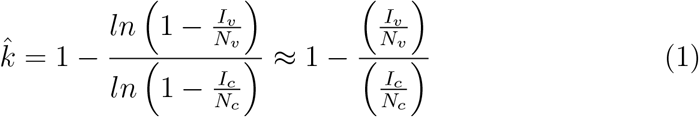

with the approximation valid for small numbers of infected compared to the trial group sizes *I/N* ≪ 1 which is well valid in all cases here. See [28] and [29], for example, for the mathematical approach used to estimate the vaccine efficacy for a dengue vaccine using the publicly available Phase III trial data.

In detail, from the initially susceptible individuals in each group one estimates the infection rate *β* from the control group, and the eventually reduced infection rate (1 − *k*)*β* in the vaccine group with efficacy *k*, via the processes

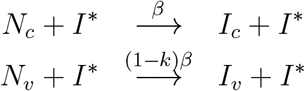

giving first a likelihood for the probability of background infection, for convenience the probability not to become infected in the control group *θ*_*β*_, i.e. *L*(*θ*_*β*_) = *p*(*I*_*c*_|*θ*_*β*_) and via the Bayesian ansatz the posterior

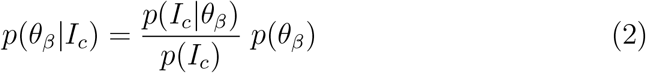

and likewise in the vaccine group, such that we finally obtain the posterior *p*(*k*| *I*_*v*_, *I*_*c*_) of the vaccine efficacy *k* only as function of the trial data (by marginalizing over the internal background infection parameter, hence the form of which is not entering into the final results) via

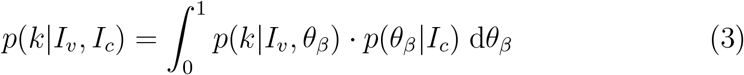

and from this also its cumulative distribution function *P* (*k*| *I*_*v*_, *I*_*c*_). In Fig. 2 a) we show the numerical results for the the Oxford/AstraZeneca vaccine efficacy posterior *p*(*k*| *I*_*v*_, *I*_*c*_), giving a good visual impression of the estimated efficacy and its insecurity due to the small trial data numbers. In Fig. 2 b) we show the cumulative distribution function *P* (*k* |*I*_*v*_, *I*_*c*_), from which one can read off the confidence intervals.

**Figure 2:**
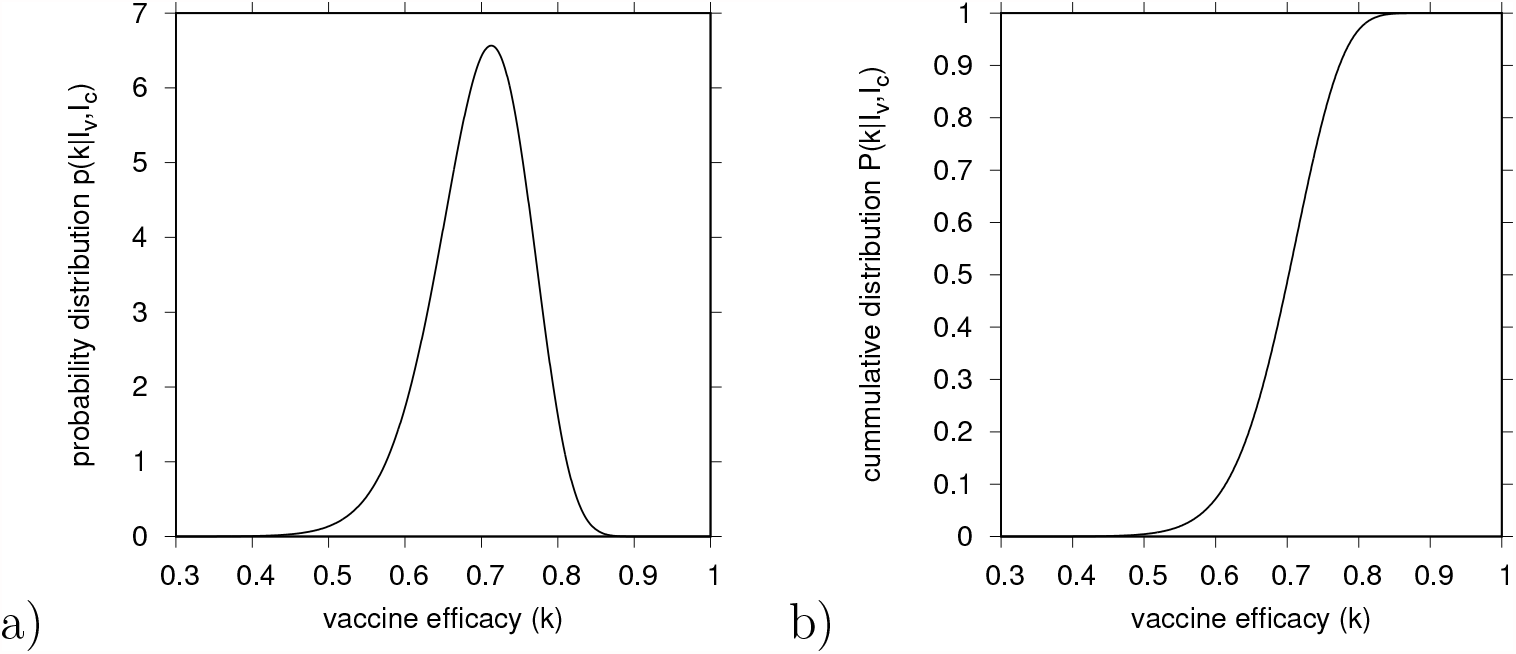
Bayesian analysis of the vaccine efficacy bases on the raw trial data of the Oxford/AstraZeneca vaccine [6]. a) Posterior distribution of the vaccine efficay *p*(*k*|*I*_*v*_, *I*_*c*_), and b) its cumlative distribution function *P* (*k*| *I*_*v*_, *I*_*c*_) to read off median and confindence intervals.

From the data which generates Fig. 2 we obtain from the median of the marginalized posterior *P* (*k*_0.5_|*I*_*v*_, *I*_*c*_) = 0.5 the Bayesian estimate of the vaccine efficacy *k*_0.5_ = 0.703 = 70.3% and the 95%-confidence interval from the 0.025 and 0.975 quantils, hence *P* (*k*_0.025_|*I*_*v*_, *I*_*c*_) = 0.025 for the lower bound *k*_0.025_ = 0.559 and *P* (*k*_0.975_| *I*_*v*_, *I*_*c*_) = 0.975 for the upper bound *k*_0.975_ = 0.805.

These estimations are in good agreement with the values given in [6] (with 70.4 % (95%-CI: 54.8 - 80.6)), with small differences due to Bayesian priors being as uninformed as possible, see [28, 29] for further details.

### 2.3. BioNTech/Pfizer vaccine: large scale population analysis

After the first period of vaccine trial data published around December 2020 (Oxford/AstraZeneca vaccine [6], BioNTech/Pfizer vaccine (mRNA vaccine) [7], Moderna vaccine (mRNA vaccine) [8], and Janssen/Johnson-&Johnson [9]), results of first larger scale population studies of vaccine efficacy became available, e.g. for the mRNA vaccine BioNTech/Pfizer [10].

In this study, data from Israel’s largest health care organization, the Clalit Health Service, were used to evaluate the effectiveness of the BioNTech/Pfizer vaccine (BNT162b2 mRNA) vaccine. Aspects of vaccine efficacy after the first dose and after the second dose are given. Such detailed studies are rare and not yet available for all the vaccines already licensed for emergency use.

For this specific study, preliminary analyses of vaccine efficacy against hospitalization and severe disease, called *k*_*H*_ in the modelling setup, and in some cases different efficacies against infection (immunizing effect of the vaccine reducing the probability of a vaccinated individual to transmit the infection), called *k*_*I*_ can be performed.

A first inspection of the given vaccine efficacies in [10] already indicates that not only the medians of *k*_*H*_ and *k*_*I*_ can be quite different, but also the confidence intervals merely overlap, or in some cases are disjunct, indicating that future studies will, most likely, not give equal efficacies against severe disease and infection. In the present case of the BioNTech/Pfizer vaccine the differences in efficacies are mostly observed after a single dose, while efficacies after the second dose are estimated to be above 90%, hence remarkably higher than for the estimated vaccine efficacy for the Oxford/AstraZeneca vaccine, for example, after the full immunization (two dose) scheme.

Data from the Clalit Health Service data were analyzed from December 20, 2020, to February 1, 2021, with all newly vaccinated persons matched in a 1:1 ratio to unvaccinated controls [10]. With *N*_*v*_ = *N*_*c*_ = 596 618 (and later keep to a good extend to equal group sizes, hence *N*_*v*_*/N*_*c*_ ≈ 1), vaccine efficacies can be obtained from the raw data given above as

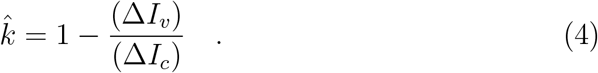

Using a refined Bayesian analysis, as described in Section 2.2, we compare the vaccine efficacy against hospitalization and against infection. First, for the one dose vaccination schedule, see Fig. 3 a), we use the data for “documented infection” with confirmed PCR with 21 to 27 day after vaccine administration. With the two distributions only slightly overlapping, a high vaccine efficacy is estimated for protection against hospitalization *k*_*H*,1_, *k*_*H*,1_ = 78%[61% − 91%], with maximum around 80%, plotted in green in Fig.3 a). Vaccine efficacy against infection *k*_*I*,1_ is estimated as *k*_*I*,1_ = 60%[53% − 66%], and has its maximum just below 60%, plotted as purple curve in Fig.3 a).

**Figure 3:**
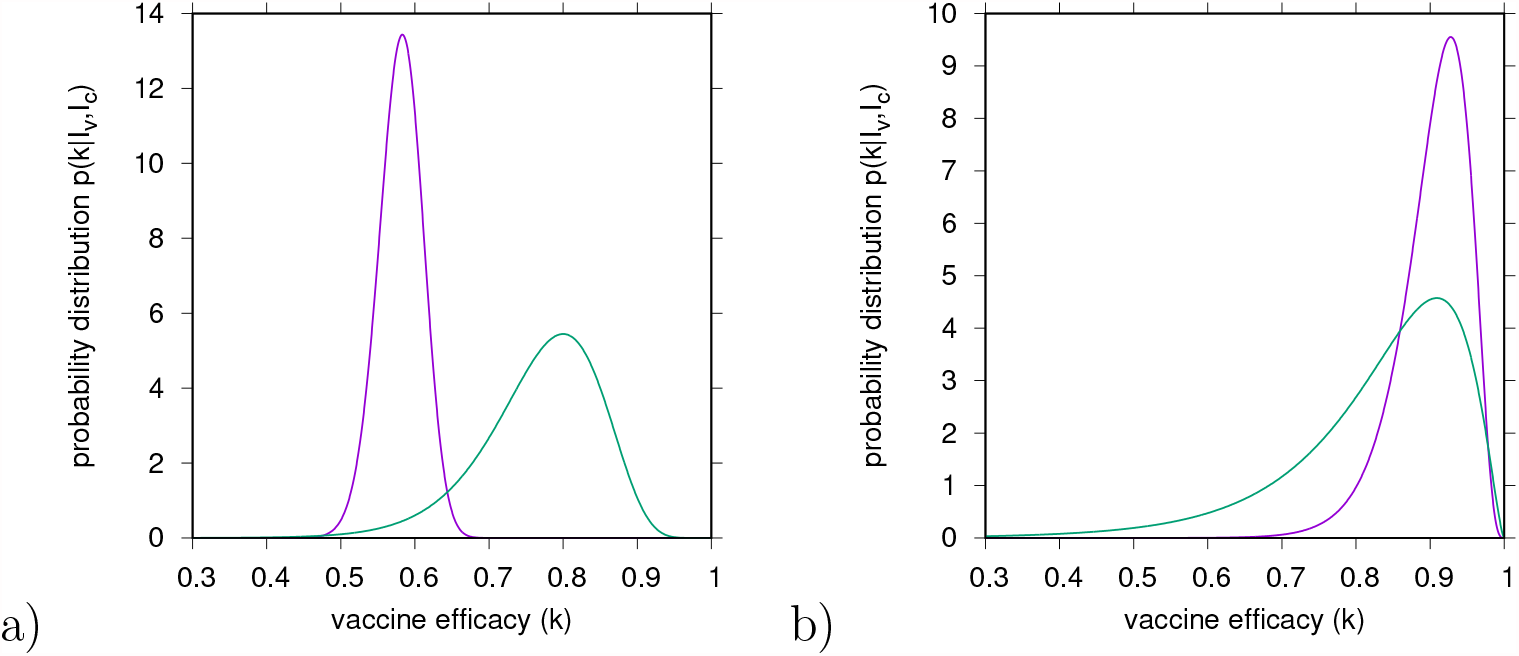
Comparison of vaccine efficacies against hospitalization *k*_*H*_, in green, and against infection *k*_*I*_, in purple. In a) vaccine efficacy is estimated for a single dose vaccine *k*_*H*,1_ and *k*_*I*,1_, for 21 to 27 days after vaccine administration and in b) vaccine efficacy is estimated for two dose vaccine *k*_*H*,2_ and *k*_*I*,2_, for 7 days after vaccine administration, the full immunization schedule to the end of the follow-up. Data were obtained from [10].

Vaccine efficacy increases significantly with the complete immunization with two dose vaccination. Here, the two distributions are well overlapping, with both, the efficacy against hospitalization *k*_*H*,2_ = 92%[88% − 95%] and efficacy against infection *k*_*I*,2_ = 87%[55% −100%], having their maxima around or above 90%, see Fig. 3 b).

The efficacy against infection is quite well measured with confidence intervals between 80% and nearly 100%, whereas the smaller trial numbers of hospitalized can leave some wider insecurity of efficacy with lower bound as far down as 50 to 60%. Nevertheless, the bulks of the distributions overlap well, and for modelling purposes we assume, here, roughly equal protection against hospitalization and against infection for the two dose vaccine, *k*_*H*_≈ *k*_*I*_≈ 92%, as given in [10]. It is important to consider the reduction of efficacies due to the new variants. New information on vaccine efficacies are released frequently and can be included in the modeling framework as needed. Moreover, please, note that the actual numbers of median efficaies are not that important on the second or third digit, due to still large confidence intervals in the studies, but the order of magnitude is informative in modelling exercises as presented here.

Nevertheless, the first dose vaccination regime is still important to be considered in the momentary scenario in which populations have large proportion of the vaccinated individuals with a single dose, still awaiting to receive the second vaccine dose. As an example, the Basque Country setting in Spain, as of June 14, 2021, see Fig. 4, counting 47.6% and 31.3% of the population vaccinated with at least one dose and with the complete immunization schedule respectively. Note that the full immunization schedule considers two dose for BioNTech/Pfizer, Moderna and Oxford/AstraZeneca and a single dose for Janssen/Johnson&Johnson. Official data on vaccination doses are updated every day 15 of each month.

**Figure 4:**
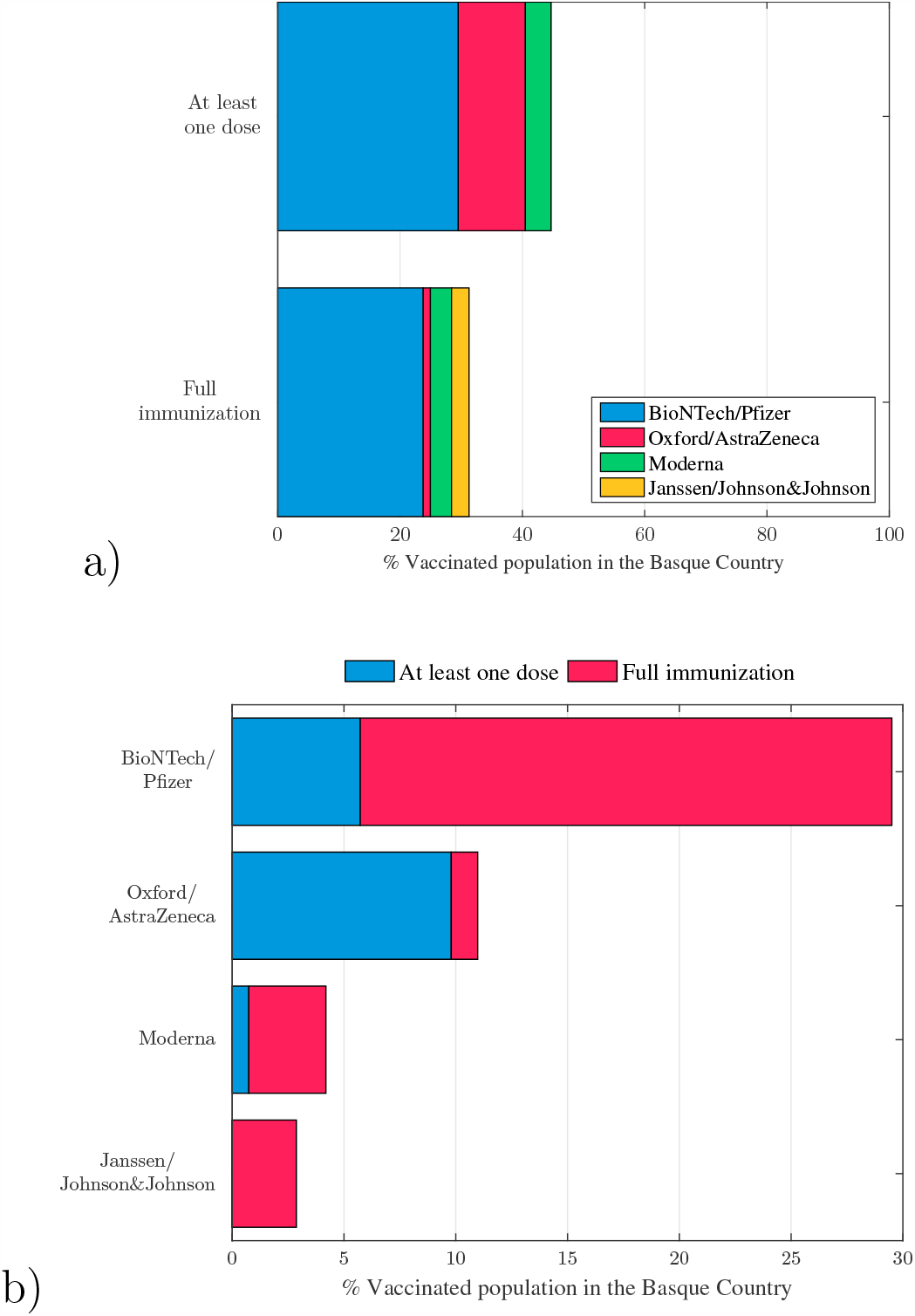
Vaccination roll out in the Basque Country, Spain, as of June 14, 2021. Four vaccines are approved for emergency use: BioNTech/Pfizer & Moderna (mRNA vaccines) and Oxford/AstraZeneca & Janssen/Johnson&Johnson (viral vector vaccines). In a) the vaccination coverage of individuals that have received at least a single dose of a vaccine (≈ 47.6%) and the coverage of individuals that have completed the immunization schedule (≈ 31.3%). In b) the detailed numbers of doses per vaccine type administered in the Basque Country, Spain, up to June 14, 2021, given in percentage.

**Figure 5:**
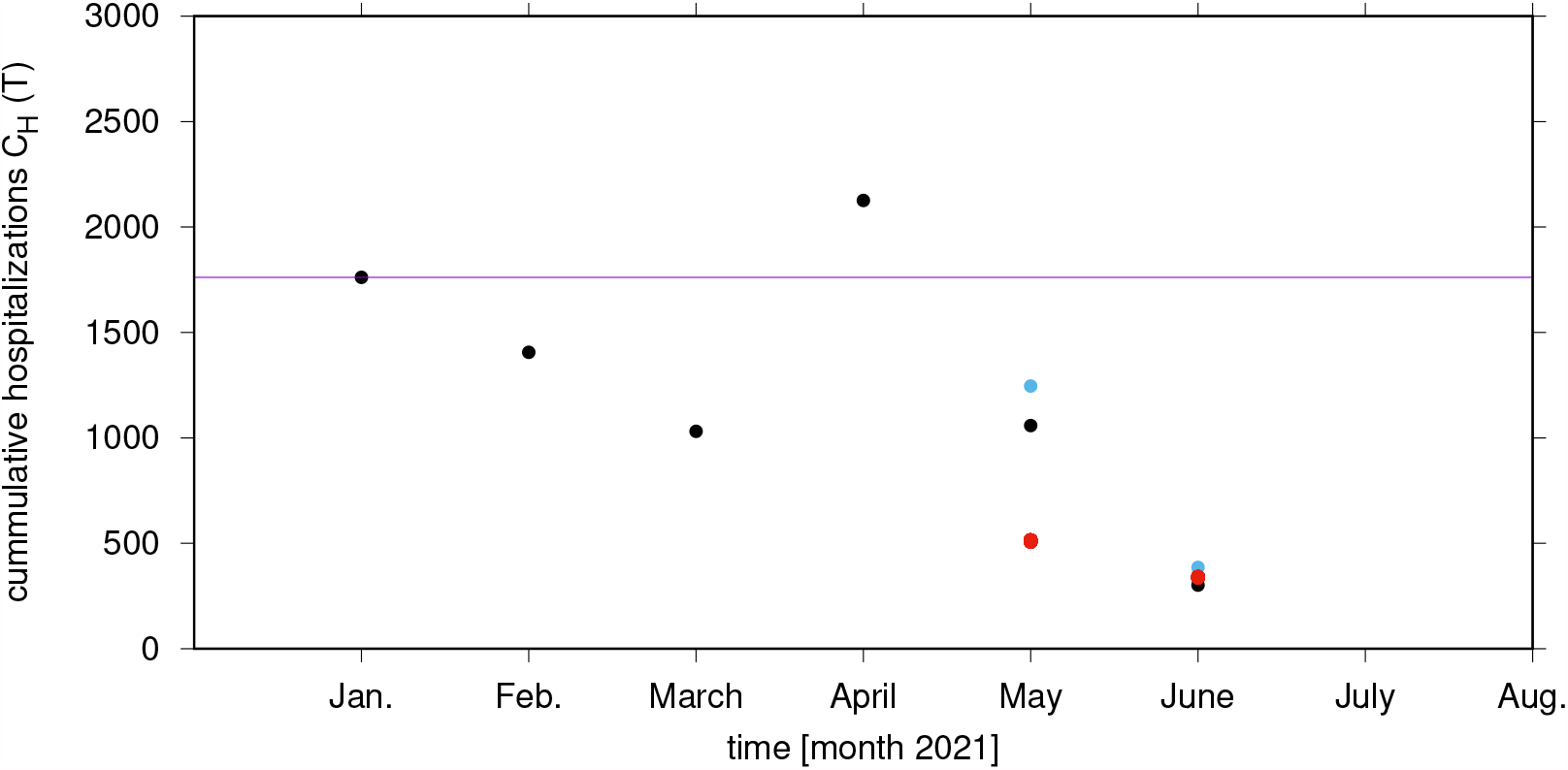
Evaluation of the vaccination impact on hospitalizations in the Basque Country for May and June 2021 (plotted as red dots) in comparison with the official data on hospitalizations in the Basque Country from January to May 2021 and with preliminary data for June, up to 29th (plotted as black dots).

## 3. Vaccination coverage to achieve herd immunity: basic concepts using the simple SIR model

Differences in vaccine efficacies after the full immunization schedule with two doses, e.g. in the compared efficacies of Oxford/AstraZeneca of roughly 70 % and BioNTech/Pfizer of above 90 %, affects the vaccination coverage needed to achieve herd immunity by vaccination.

Here, we present the basic concepts of a simple SIR model, which are already quite informative, and later refine our modelling framework to evaluate the current situation in the Basque Country, Spain, and other European regions with a mixed vaccination coverage of first and second dose, and eventually mixed vaccine efficacies against hospitalization and infection.

From a simple SIR model with

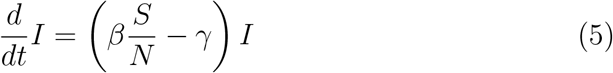

and the vaccination coverage *c* of population *N* as *c* ·*N*, hence remaining susceptible individuals *S* = (1−*c*)*N*, we use the condition of zero growth *λ* = 0 as threshold condition for the vaccination coverage *c* via

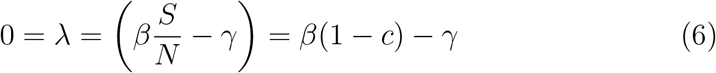

giving

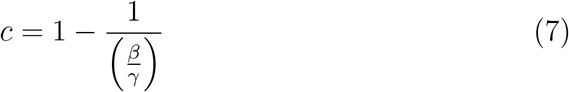

as threshold coverage to obtain the population herd immunity *λ* ≤ 0. This is the classically used formula *c* = 1 − 1*/*ℛ_0_ for vaccination coverage threshold in function of the basic reproduction ratio ℛ_0_.

With a perfect vaccine *k* = 1, the herd immunity threshold is driven by the so called ℛ_0_ of a disease. As an example, for *β* ≈ 3.5*γ* (an estimated ℛ ℛ_0_ = 3.5), we obtain

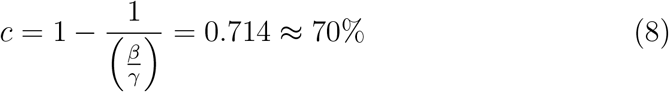

vaccine coverage, with full amount of doses as aim to obtain herd immunity in the population. This value has being frequently mentioned in the public media during the COVID-19 pandemic.

However, in real life, the situation is more complex, since vaccines are imperfect *k <* 1 and therefore herd immunity by vaccination depends not only on the ℛ_0_ value but also on the given vaccine efficacy of the vaccines administrated in the population. Moreover, applied to COVID-19, the already gained immunity via natural infection will eventually play a role for the population coverage needed to achieve the herd immunity status.

Some useful contributions in this direction are provided by [30, 31], for example, where models for SARS-CoV-2 considering heterogeneity on the population level or overall vaccine efficacy is considered. However, to our knowledge, this is the first exercise considering heterogeneity on vaccine efficacy for a single dose versus two dose immunization schedule, including population immunity by natural infection.

As such, we will present in the next Section, a refined model to include variability between vaccine efficacies after a first dose and after a second dose. With this modeling framework the eventual differences in protection against severe disease and against infection for different vaccines can be also evaluated, as soon as empirical evidence such as in [10] becomes available.

### 3.1. Considerations for imperfect vaccines and population immunity by natural infection: the Basque Country as a case study

Applied to the current epidemiological scenario in the Basque Country, Spain, with vaccination roll out using imperfect vaccines, we consider empirical vaccines efficacy *k <* 1 and a proportion of already naturally immunized persons via previous natural COVID-19 infection as recorded by the public health managers. With a population size of *N* = 2.2· 10^6^, less than 200 000 infections were reported as of July 1st 2021, around 10% of the population been already immune prior to vaccination, a pool of default susceptible individuals *S*_0_*/N* ≈ 90% is considered. This assumption can be modified as new positive cases are detected to be included in the analysis counting the current immunized population via natural infection or by vaccination at a given time. Vaccination coverage is given by *cN* = *c*(*S*_0_ + *R*(*t*_0_)) = *cS*_0_ + *cR*(*t*_0_) with the recovered *R*(*t*_0_) at a given time *t*_0_of analysis, when vaccines are administrated in the population independently of the individual previous record of negative or positive PCR test.

For non-vaccinated (1 − *c*)*S*_0_ and vaccinated with vaccine efficacy *k*, i.e. *c*·(1 −*k*)*S*_0_, where *r* = 1 −*k* is the relative risk measured in vaccine trials, we obtain now a refined version of the dynamics of infected

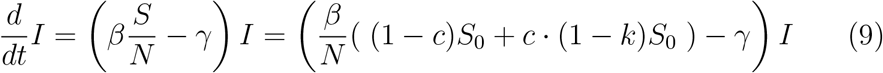

giving via the growth condition of *λ* = 0 the result for the vaccination coverage threshold

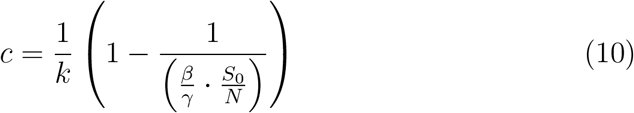

showing that 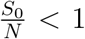 reduces the coverage threshold, but *k <* 1 can significantly increase the threshold again.

Here, as an example, as shown in Section 3, we consider an infection rate of

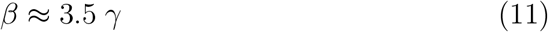

and a proportion of the susceptible population as

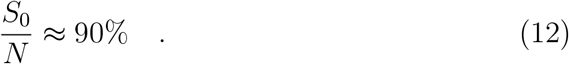

By assuming vaccination roll out with a perfect efficacy *k* = 1, the value of vaccination coverage to achieve herd immunity is given by

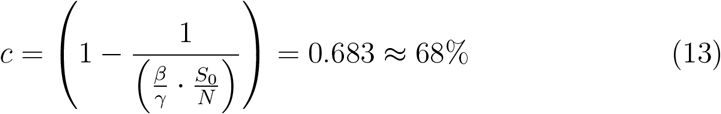

which is only slightly below the value estimated for a 100% susceptible population, i.e, considering no acquired natural immunity, of *c* = 0.714 ≈71%.

On the other hand, by assuming vaccination roll out with an imperfect efficacy *k <* 1, such as estimated in Section 2.2 and in Section 2.3, the vaccine efficacies to be assumed here are, for example, in the case of the Oxford/AstraZeneca vaccine with an estimated efficacy of

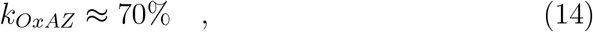

and for the BioNTech/Pfizer vaccine, with a reported efficacy of

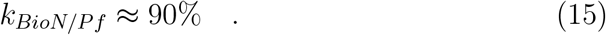

The results for the vaccination coverage threshold to obtain herd immunity are different, assuming vaccination roll out with one or another vaccine.

When assuming that the Basque population will receive only the BioN-Tech/Pfizer vaccine, we obtain the following result for the vaccination coverage threshold to reach population herd immunity

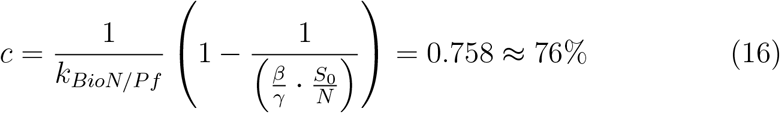

which is close to what is expected for perfect vaccine efficacy. Here, the vaccination coverage of *c* = 0.714 ≈ 71% for vaccine efficacy of *k*_*BioN/P f*_ ≈ 95% is extremely good as in a scenario of a perfect vaccine protecting again overall infection.

On the other hand, for the example of the Oxford/AstraZeneca vaccine with *k*_*OxAZ*_ ≈ 70% efficacy we obtain

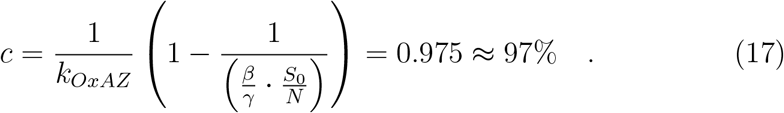

for the vaccination coverage threshold to reach the population herd immunity.

This result is surprising and must be considered carefully in the case of using the vaccine with significantly lower efficacy only, needing a very high vaccination coverage of more that 95% to achieve herd immunity.

In the next Section we will describe a further refined modelling framework to include various aspects of the variability of the vaccines, considering different efficacies as well as their performance against severe disease and against overall infection.

## 4. Modeling COVID-19 with the basic SHAR model

Applied to COVID-19 pandemic in which mild and severe cases of infection are well distinguished, we extend the basic SIR (Susceptible-Infected-Recovered) modeling framework into the so-called SHAR model [14], where the infected class is stratified into Hospitalized/severe disease cases (*H*) and Asymptomatic/mild cases (*A*). With infection rate *β* and recovery rate *γ*, susceptible *S* individuals becoming infected can either develop severe disease prone to hospitalization *H*, with a proportion *η*, or develop a mild infection, potentially asymptomatic *A*, with a proportion (1 −*η*). A scaling factor *ϕ* is used to differentiate the infectivity *ϕβ* of mild/asymptomatic infections in respect to the baseline infectivity *β* of severe/hospitalized cases. The value of *ϕ* can be tuned to reflect different situations. While a value of *ϕ <* 1 reflects the fact that severe cases have larger infectivity than mild cases (e.g., enhanced cough and sneeze), a *ϕ >* 1 value indicates that asymptomatic individuals and mild cases contribute more than severe cases to the spread of the infection (e.g., due to their higher mobility and contacts). Recovered individuals *R* are considered resistant to reinfection. The dynamics for the mean values can be written as ordinary differential equation system

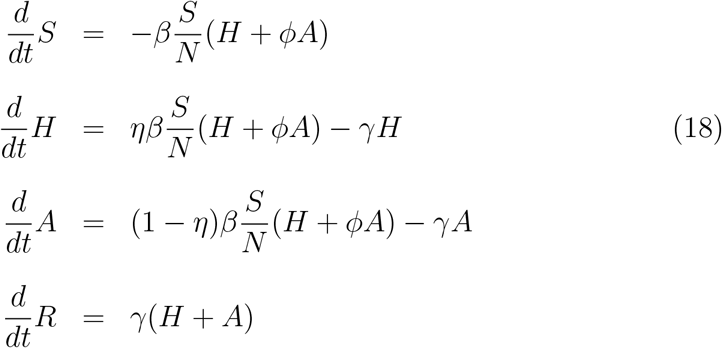

and the vaccination coverage threshod to achieve herd immunity is obtained similarly as presented for the simple SIR model in Section 3.1.

The SHAR model was extended to a *SHARUCD* modeling framework to describe the epidemiological situation of COVID-19 in the Basque Country, validated with empirical data and it is, up to date, used by the local public health managers to monitor the impact of lockdown measures [14, 15, 16, 17, 18, 19].

### 4.1. Modeling vaccine efficacy against severe disease or/and infection: the SHARV

A vaccine which protects against severe disease but not against infection needs to be modelled in a SHAR framework to distinguish the remaining risk *r* = 1−*k* for hospitalization and no effect at all against mild/asymptomatic infection. In the last case, the undetected cases will contribute to the force of infection. Here, we distinguish naive susceptibles *S* and vaccinated sus-ceptibles *S*_*v*_, where the naive susceptibles have the natural infection rate *β* and the vaccinated the reduced infection rate (1 − *k*) · *β*, as described in the vaccine trial analysis above. Hence we have the complete model given by

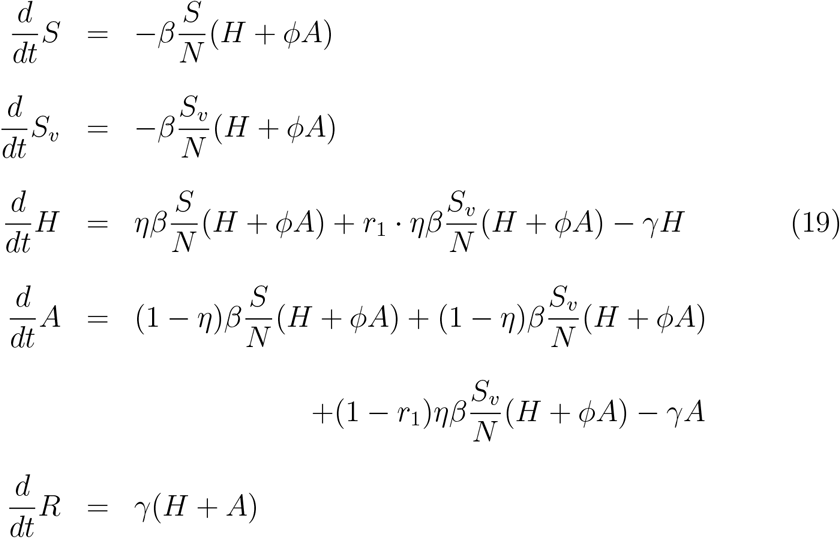

which we call a model for a vaccine of type 1, protecting against severe disease but not against infection.

On the other hand, a model for a vaccine which protects as much against infection as against severe disease is given by the following dynamical system

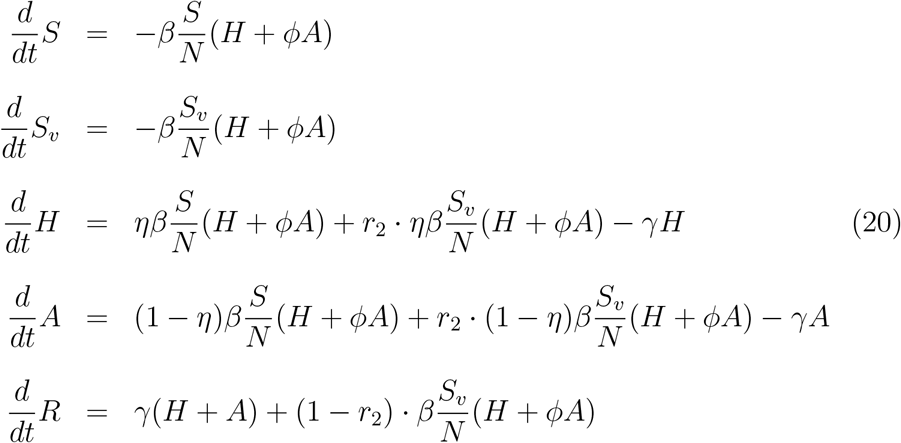

and is in its dynamical behaviour much closer to the simple SIR model as described before. We call this a vaccine of type 2.

The different COVID-19 vaccines currently in use have features placing them closer to one or the other of these two extreme cases, and as shown in previous analysis of the large population studies, real life vaccines would have aspects of the vaccine type 1, protecting against severe disease but failing to block transmission, but also some protection against infection, vaccine type 2. The differences between these vaccine types can be analyzed in detail (please see [32]), revealing a significant sensitivity to the difference of infection rate for severe cases *H* and for mild/asymptomatic *A*, which are parametrized in the SHAR modelling framework by the parameter *ϕ*.

These two limiting cases of vaccine type 1 and vaccine type 2 are refined to consider a mixed vaccination roll out scenario with different vaccines efficacy and its effects observed with a single dose versus a two dose immunization scheme. It is important to mention that there is not much reported on varying efficacies per dose administrated for the Oxford/AstraZeneca vaccine, or any other vaccine already licensed for emergency use. In this study we consider the available information for the BioNTech/Pfizer vaccine [10], assuming that the variations reported eventually capture many aspects of heterogeneous vaccine coverage and efficacies.

## 5. The SHARV model for a mixed immunization schedule: single dose and second dose vaccination roll out

To evaluate the impact of the current vaccination programmes we extend and refine the SHARV model described above. Susceptible population are now stratified into unvaccinated susceptible (*S*), susceptible vaccinated with a single vaccine dose (*S*_*v*1_) and susceptible fully immunized with two vaccine doses (*S*_*v*2_).

As reported in [10], a single vaccine dose has a significant lower efficacy as compared to the full immunization schedule with two vaccine dose. Furthermore, it has been reported that in a single dose regime the efficacy against hospitalization and severe disease is significantly higher than the efficacy against infection, i.e. the so called sterilizing immunity, occurring when vaccinated individuals cannot transmit the virus.

These differences are taking into account and the model framework is flexible enough to also consider different vaccines, different vaccine efficacies and different vaccine coverages. Vaccines performance are reported in term of relative risk *r* = 1− *k* and vaccine efficacy *k*. Vaccination is implemented by assuming reduced infectivity *r*· *β* (for vaccinated group) against natural infectivity *β* (for non-vaccinated group), as measured in vaccine trials and described above.

In this model, different vaccines with various efficacies against severity and against infection can be evaluated. By taking different efficacies into account after first dose *k*_*H*,1_ and *k*_*I*,1_, and eventually second dose *k*_*H*,2_ and *k*_*I*,2_, hence *k*_*H,j*_ and *k*_*I,j*_ for *j* ∈ {1, 2}, labeling the first and second dose of vaccine, the dynamical system of the model is given by

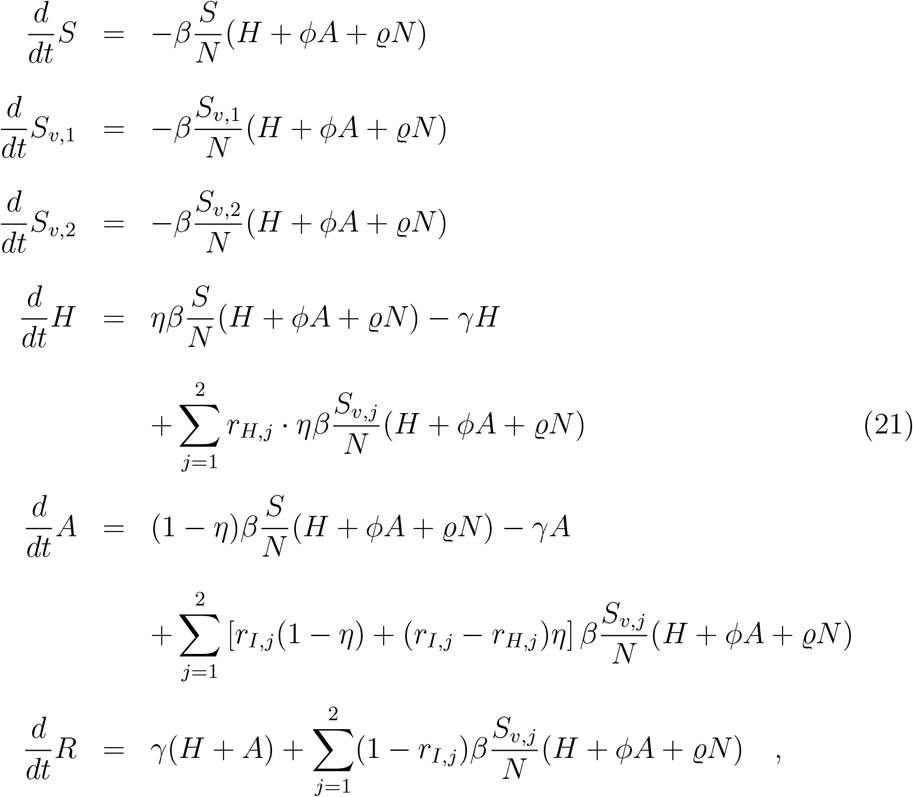

with vaccine coverage *S*_*v*,1_ = *c*_1_*S*_0_ for first dose uptake, *S*_*v*,2_ = *c*_2_*S*_0_ for second dose uptake, and finally *S* = (1 − (*c*_1_ + *c*_2_))*S*_0_ for non vaccinated susceptible individuals, with *S*_0_ = *N* − *R*(*t*_0_).

### 5.1. Analytical solutions and numerical experiments

For analytical insights into the behaviour of the model with a single dose vaccination and two dose vaccination compared to the non-vaccination scenario, we consider the dynamics of the disease compartments

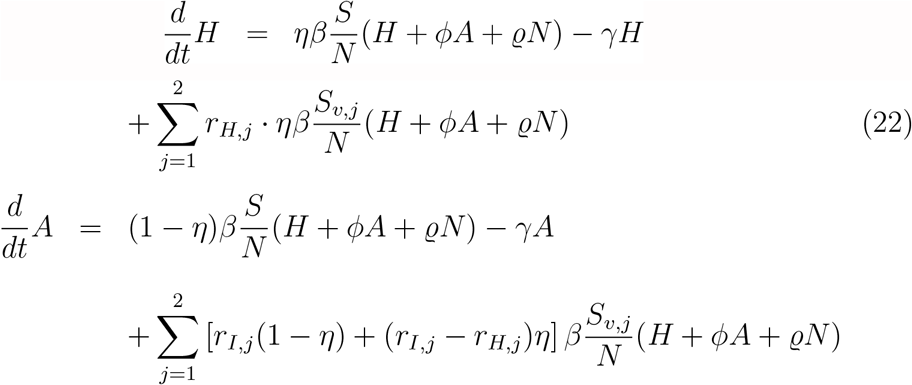

including the vaccination coverage vector *<u>c</u>* := (*c*_1_, *c*_2_) for single dose and two dose vaccine administration as the fraction of vaccinated susceptible individuals over the total number of susceptible *S*_0_ := *S* + *S*_*v*1_ + *S*_*v*2_, hence *S*_*v*1_ = *c*_1_·*S*_0_, *S*_*v*2_ = *c*_2_ ·*S*_0_ and the naive susceptible *S* = 1−(*c*_1_+*c*_2_)·*S*_0_ and for vaccine efficacy vector *<u>k</u>* = (*k*_*H*,1_, *k*_*I*,1_, *k*_*H*,2_, *k*_*I*,2_) for the respective efficacies against hospitalization and against infection after administering one dose or two doses.

### 5.2. Stationary state solutions and relative hospitalization reduction

Our model considers an imperfect vaccine with vaccinated individuals able to transmit the disease even when the vaccine is reported with a significant efficacy. Moreover, we assume the COVID-19 herd immunity is not yet reached (please see Section 3.1), and therefore, we consider *S*_0_(*t*_0_) =: *N*− *R*(*t*_0_). The stationary state solution is

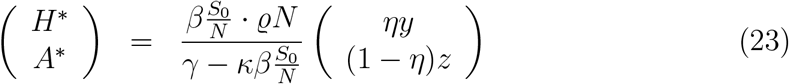

now with vaccination specific variables

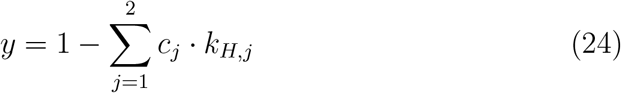

and

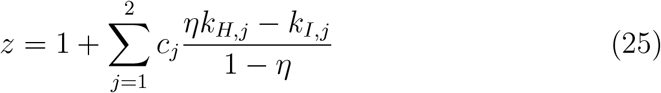

using *κ* := *ηy* +*ϕ*(1− *η*)*z* = *κ*(*<u>c</u>, <u>k</u>*) in the stationary state solution, depending on the vaccination variables *y, z* and on the SHAR specific parameters *η* and *ϕ*.

For the relative reduction of hospitalizations we have

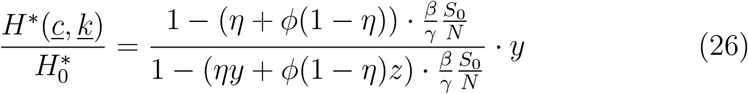

with vectors for vaccination coverage *<u>c</u>* = (*c*_1_, *c*_2_) for one dose and two doses respectively, and for vaccine efficacies *<u>k</u>* = (*k*_*H*,1_, *k*_*I*,1_, *k*_*H*,2_, *k*_*I*,2_) for the respective efficacies against hospitalization and against infection for one dose or for two doses.

## 6. Vaccination impact in the Basque Country

To investigate the vaccination impact in the Basque Country, Spain, as a first conceptual study taking variable vaccine efficacies and vaccine coverages into account we use the values obtained in [10], as presented in Section 2.3. We evaluate the vaccine impact on severe cases/hospitalizations of COVID-19 in the Basque Country, Spain, assuming the reported vaccine coverages, including the present population status of remaining suceptibles after one year of natural infection, and considering vaccine efficacies as reported from Israel in, [10].

First, to make a projection of the vaccine impact on hospitalization for the end of May 2021, vaccine efficacy vector is given by *<u>k</u>* = (*k*_*H*,1_ = 78%, *k*_*I*,1_ = 60%, *k*_*H*,2_ = 92%, *k*_*I*,2_ = 92%) and from the vaccination coverage, as of 23 May 2021, of *c*_2_ = 17.5% for coverage of two doses vaccinated, and (*c*_1_ +*c*_2_) = 38.8%, hence *c*_1_ = 21.3% for single dose coverage alone, giving the vaccination coverage vector *<u>c</u>* = (*c*_1_ = 21.3%, *c*_2_ = 17.5%), we obtain the vaccination related variables to be

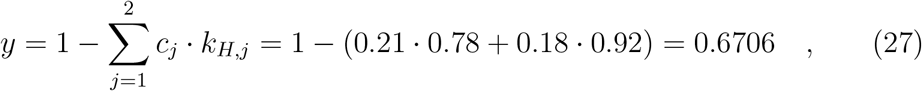

and by assuming *η* = 0.08 = 8% for the hospitalization ratio in the Basque Country,

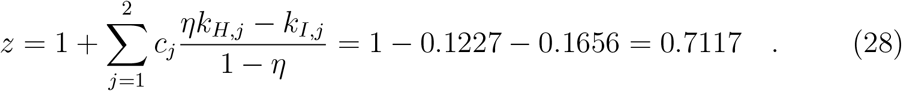

Further, with around 200 000 identified infected cases over the past year, representing roughly the number of recovered individuals from COVID-19 in the Basque Country,

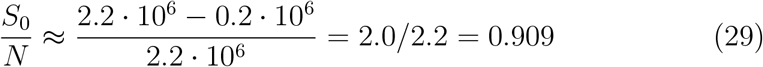

gives a first approximation for the remaining suceptibilty in the study population.

Results for the evaluation of the vaccination impact on hospitalizations in the Basque Country are presented in Fig. 6. When considering the number of hospitalizations in January 2021 as baseline, assumed to have no significant effects on population immunity by vaccination that only started in the end of December 2020, we observe first a significant reduction of hospitalizations with present vaccination uptake at mid June (see red and black dots in month 6, June, in Fig. 6), whereas in May the expected reduction was not reached at all, and with the April value above expectations, probably due to large subcritical fluctuations [18, 19], see Fig. 6, plotted in black in month 4, April.

In detail, we first obtain the overall hospitalizations for each month of 2021, from January 1st onward (black dots). Data was continually updated.

The value available at 15th of May was multiplied by two to obtain an approximation for the total hospitalizations notified for the whole month (blue dot at month 5, May, in Fig. 6). This is a reasonable assumption as the current setting is shown to be in quasi-stationarity with large fluctuations, see [18, 19]. The expected number of hospitalizations from the reduction 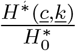 to baseline in January, including a proportionality of reported incidence with the calculated prevalence’s

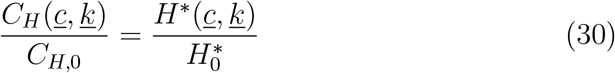

which gives an evaluation of the vaccination impact from *C*_*H*,0_ = *C*_*H*_ (*T* = Jan 2021) without vaccination as baseline to the values in May 2021 as

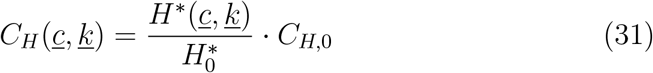

with the vaccination coverage from May 23, 2021, as *<u>c</u>* = (*c*_1_ = 21.3%, *c*_2_ = 17.5%), shown as red dot at month 5 in Fig.6.

This procedure gave still unsatisfactory results for May 2021, due to the large fluctuations observed also between January and April, see [18, 19] for further discussion on such large subcritical fluctuations and import. When repeating this exercise in mid of June, now with updated two doses coverage as 34.1% and with at least one dose coverage as 50.9%, we have *<u>c</u>*_*n*+1_ = (*c*_1_ = 16.8%, *c*_2_ = 34.1%) for June, as opposed to the May values of *<u>c</u>*_*n*_ = (*c*_1_ = 21.3%, *c*_2_ = 17.5%). We then projected from the 15th of June value for hospitalizations the full number expected for end June (blue dot at month 6 in Fig.6). The data point available for the 29th of June (black dot at month 6, June of 2021, in Fig.6) is still lower than the expected value, however, much closer than observed for May, with the expected vaccination impact on the hospitalization ratio (red dots) agreeing qualitatively well with the empirical data (black dots).

## 7. Discussion and conclusions

We have analysed the impact of variable vaccine efficacies and coverages on the reduction of severe disease/hospitalization of COVID-19 in an example of the Basque Country epidemiological setting. To evaluate the vaccine effects on sever cases, a mathematical modeling framework considering heterogeneity on vaccine efficacy for hospitalization and overall infection, including population immunity by natural infection, was developed and analyzed.

We use the recent results of vaccine efficacies from large scale population surveys and although we have considered simplified assumptions for the remaining levels of susceptibles and the efficacies for mainly one vaccine, results are consistent with the presently available data, since this vaccine accounts for the majority of vaccinated individuals in the Basque country. However, it is important to mention that there is still an ample space for further evaluations, including additional stochastic effects as described e.g. in [18, 19, 33] and appearing also in the large confidence intervals so far observed in the vaccine efficacies in vaccine trials as well as in larger population studies, see e.g. [6, 10] representative for other such studies recently published.

Information on COVID-19 vaccine efficacies are updated frequently and the new information can be included into the modeling framework as needed. Studies like the one described here are, nevertheless, timely and of major importance to understand the vaccination coverage needed to achieve herd immunity in different settings.

Differences of vaccine efficacy against severe disease versus vaccine efficacy against overall infection after the full two dose immunization regime in the uneven vaccination roll our settings are the goal for understanding the real impact of COVID-19 vaccines worldwide, using different types of vaccines at the same time, i.e., mixed efficacies of two different vaccine classes.

For that, extensions to evaluate a single dose versus two dose immunization schedule efficacy are also included in the minimalistic SHARV framework which is, nevetheless, the baseline model to evaluate different scenarios when new empirical evidence for COVID-19 vaccines performance becomes available.

Insights on how to best combine the use of the available COVID-19 vaccines optimizing the reduction of hospitalizations are of major interest, since the existing vaccines are imperfect, leaving a proportion of the population at risk of acquiring the infection and eventually developing severe disease. Although the vaccination roll out are advancing fast, large part of the population are still covered with a single dose of different vaccines. Finally, the results presented here give a robust conceptual framework to evaluate vaccines impact not only for the next few months, when eventually the herd immunity threshold might be achieved, but also to evaluate new vaccine generations in the case of vaccine waning immunity or even when immune escape by new variants might be observed, leading eventually also to reinfection[34] and with further evolutionary effects of large fluctuations [33].

Finally, it is important to pint out that further monitoring will be needed since other factors such seasonality of respiratory diseases might play an additional role on disease transmission and control.

## Data Availability

Data is available upon request.

## Acknowledgments

This work was supported by the Basque Government through the “Mathematical Modeling Applied to Health” Project, BERC 2018-2021 program and by Spanish Ministry of Sciences, Innovation and Universities: BCAM Severo Ochoa accreditation SEV-2017-0718. M. A. has received funding from the European Union’s Horizon 2020 research and innovation programme under the Marie Skłodowska-Curie grant agreement No 792494, during which part of the present work has been conceived and started to be developed.

## CRediT authorship contribution statement

Maíra Aguiar and Nico Stollenwerk: Conceptualization, Methodology, Numerical Simulations, Writing-Original draft preparation. Joseba Bidaurrazaga Van-Dierdonck, Javier Mar, Oliver Ibarrondo and Carlo Estadilla: Data Curation, Reviewing and Editing. All authors have read and agreed to the published version of the manuscript.

